# Human Brucellosis in Iraq: what does a spatiotemporal data analysis from 2007-2018 reveal?

**DOI:** 10.1101/2023.11.13.23298496

**Authors:** Ali Hazim Mustafa, Hanan Abdulghafoor Khaleel, Faris Al-Lami

## Abstract

**Background:** Brucellosis is both endemic and enzootic in Iraq, resulting in long-term morbidity for humans as well as economic loss. No previous study of the spatial and temporal patterns of brucellosis in Iraq was done to identify potential clustering of cases.

**Objectives:** This study aims to detect the spatial and temporal distribution of human brucellosis in Iraq and identify any changes that occurred from 2007 to 2018.

**Methods:** A descriptive cross-sectional study using secondary data from the Communicable Diseases Control Center / Ministry of Health surveillance section. We used Moran’s I, local Getis-Ord’s Gi*, and the local indicators of spatial association (LISA) to detect the spatial distribution of the data. The data was analyzed using Excel software and the Quantum GIS (QGIS) version 2.18.20 (Steiniger and Hunter, 2013).

**Results:** 50,621 human brucellosis cases were reported during the 12-year study period. Human brucellosis persisted annually in Iraq across the study period with no specific temporal clustering of cases. In contrast, spatial clustering was predominant in the Northern region of Iraq.

**Conclusion:** There were significant differences in the geographic distribution of brucellosis. The number of cases is highest in the north and northeast of the country, which has borders with nearby countries. In addition, people in these areas depend more on locally made dairy products, which can be inadequately pasteurized. Despite the lack of significant temporal clustering of cases, the highest number of cases were reported during Summer and Spring.

**Recommendations:** Considering these patterns when allocating resources to combat this disease, determining public health priorities, and planning prevention and control strategies is important.

## Introduction

Brucellosis is one of the most widely spread zoonotic diseases in the world, responsible for enormous economic losses and considerable human morbidity in endemic areas.^1^ WHO estimates 500,000 annual new infections in over 170 countries, and 9% of them were from the Eastern Mediterranean region.^2,3^

Most human infections are subclinical (90.0%), resulting in delayed diagnosis and complications.^1^ Human-to-human transmission is rare, and human infection can result from direct contact with infected animals or their products, consumption of contaminated raw milk and milk products, or inhalation. The main sources of brucellosis infection are most goats and sheep (*Brucella melitensis*), cattle(*Brucella abortus*), pigs (*Brucella suis*), and dogs (*Brucella canis*) (dogs).^3^ Brucellosis seasonality shows no specific pattern. However, it usually coincides with the livestock breeding season, spring.^3^ Human exposure to livestock or their contaminated products will occur during spring; therefore, more human infections will occur where infected livestock are.^4^

A study from northern Iraq showed that the prevalence of brucellosis in livestock varied from 1% to 70%, depending on the species and diagnostic methods.^4^ Veterinary vaccination program started in 2007. However, its implementation was negatively affected by insecurities in the region.^4^

By now, no satisfactory vaccine is available for humans. ^**(9)**^ Brucellosis control depends on testing and isolating/slaughtering positive animals, vaccinating susceptible animals, and controlling animal movements ^**(10)**^.

In Iraq, there is no prior description of the spatiotemporal epidemiology of human brucellosis. This study uses MOH’s official data to identify potential changes in the spatial and temporal occurrence of human brucellosis cases in Iraq during 2007-2018.

## Methodology

### Study design

This is a descriptive retrospective study of brucellosis spatial and temporal distribution from 2007 to 2018.

### Data source

Brucellosis data were extracted from the surveillance database at the Surveillance Section at the Communicable Diseases Control Center, Public Health Directorate, Ministry of Health in Iraq. The data included the number of cases classified by the reporting districts for all 18 provinces in the country. The information data for the study include gender, age group, the reporting district, and time of diagnosis (year). The number of brucellosis cases in Iraq during 2007-2012 was retrieved from Iraq CDC as a total number of patients at the provincial level. From 2012 to 2018, the data was aggregated by province, district, age group, and sex.

Up to 2018, all 18 Iraqi governorates must report cases of brucellosis to the surveillance section monthly. The presumptive diagnosis was made by the Rapid Brucellosis Test (RBT) test, while the confirmatory test was done using ELISA and/or PCR test.

Data of each governorate’s total population in Iraq and the population distribution by age and sex were retrieved from the Central Statistical Organization CSO/ Ministry of Planning/ Iraq/ and used to calculate the incidence for the years (2007-2018).

No data about the population in each district of Kurdistan provinces for 2007, 2008, and 2009 were available in the CSO. Therefore, data from 2007-2012 were analyzed in graphs and tables using Excel software 2019. The 12-year study period was grouped into two-year periods, one from 2007-2012 and two from 2013-2018, to analyze and describe the spatial and temporal distribution of the disease.

#### Statistical analysis

A descriptive analysis of HB during period one was done by calculating the cases’ frequency and percentage according to age group, sex, and season. In addition, we described the trend of sex and age group distribution from 2007 to 2018 using staked 100% bar charts to detect any changes during this period.

We used Local Getis-Ord’s Gi* to identify whether the localized concentration is the low or high attribute zone. We used the Getis-Ord Gi* statistic to identify clusters of high magnitude. Maps and tests were done using GeoDa software V 1.12.1.161 September 2018.

Administrative and ethical approval was granted from the Public Health Directorate/Iraq MOH.

## Results

The total number of human brucellosis cases reported in Iraq from 2007 to 2018 was 51,508. The disease trend showed two peaks, one in 2008 and another in 2010, with recorded cases of 7,399 and 7,064, respectively. There was an apparent decline in reported cases from 2013 to 2018, as shown in Figure (1).

**Figure 1:**
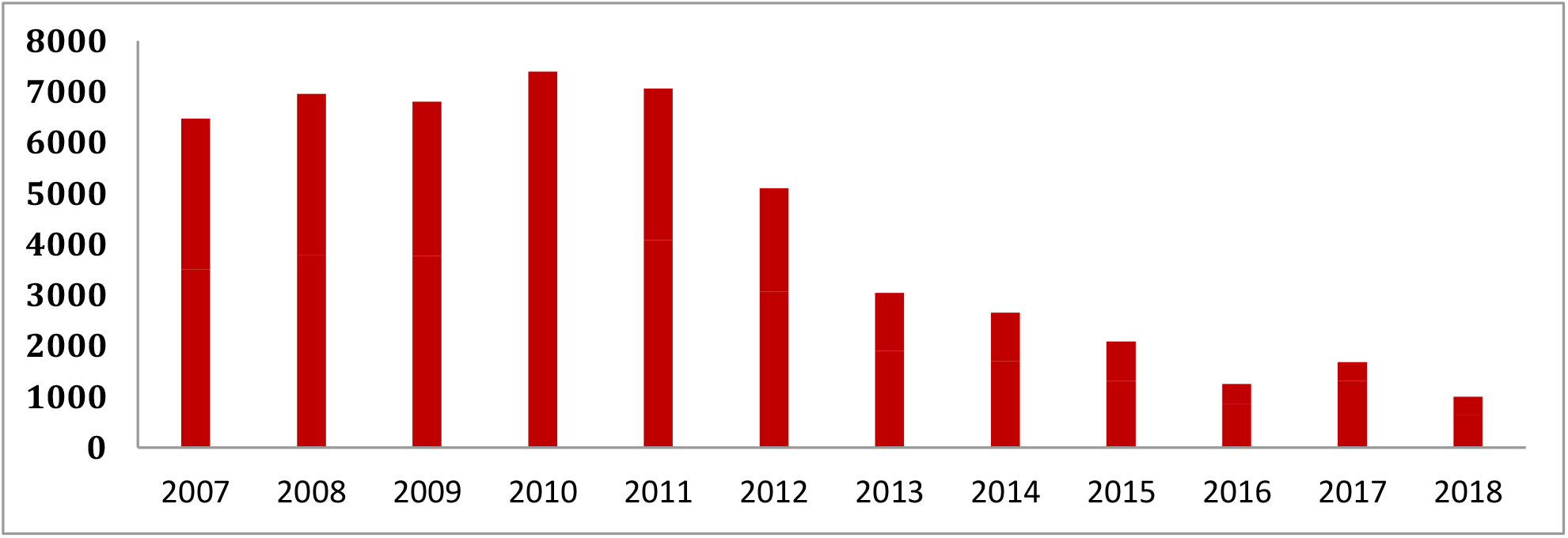
Frequency of reported cases of brucellosis in Iraq from 2007 to 2018.

**Figure 2:**
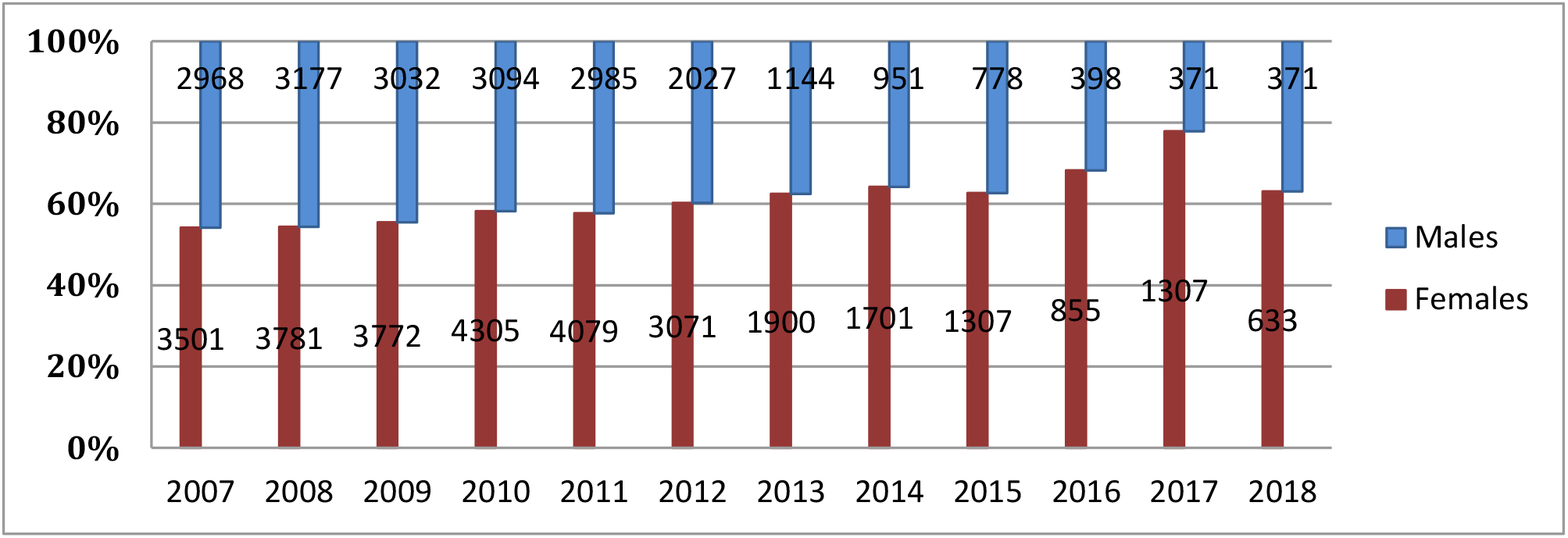
Sex distribution of human brucellosis cases in Iraq, 2007-2018.

**Figure 3:**
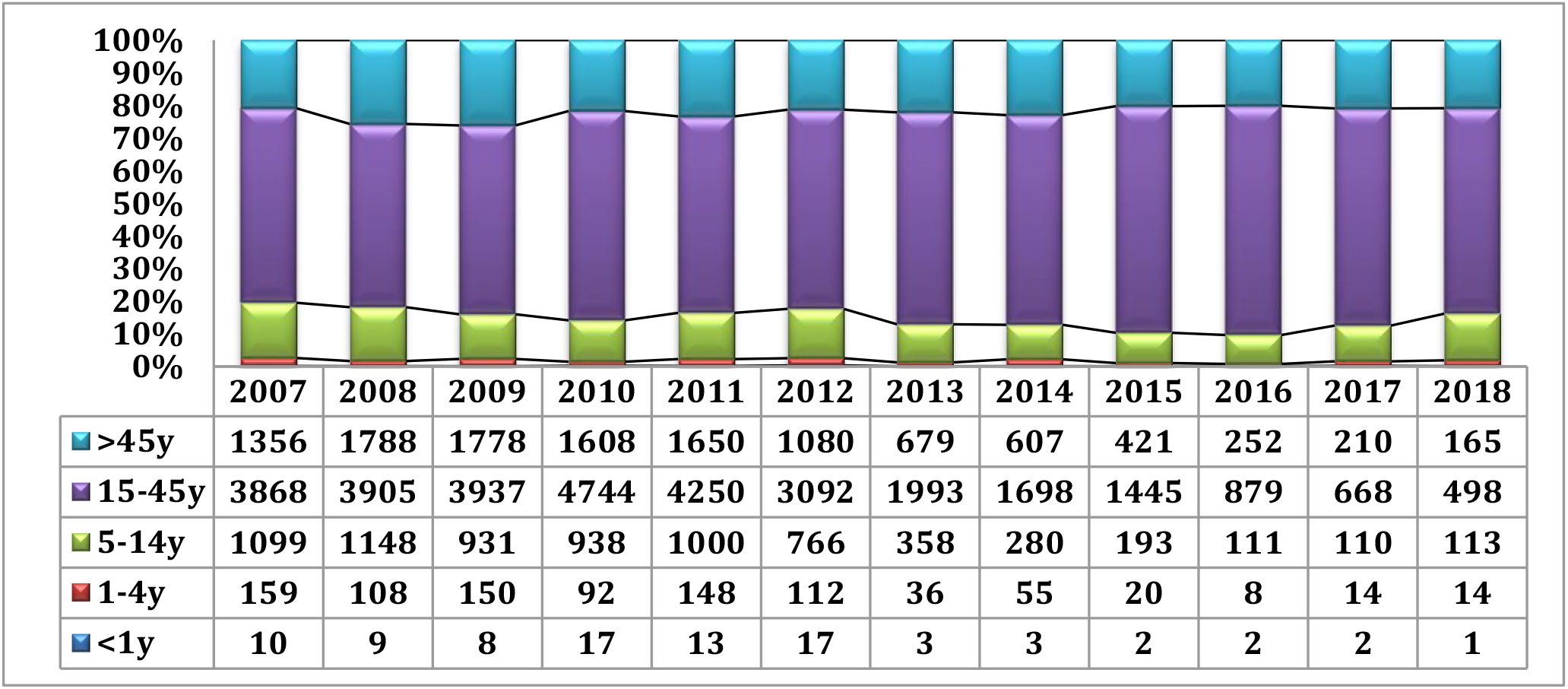
Age group distribution of human brucellosis cases in Iraq, 2007-2018.

**Figure 4.**
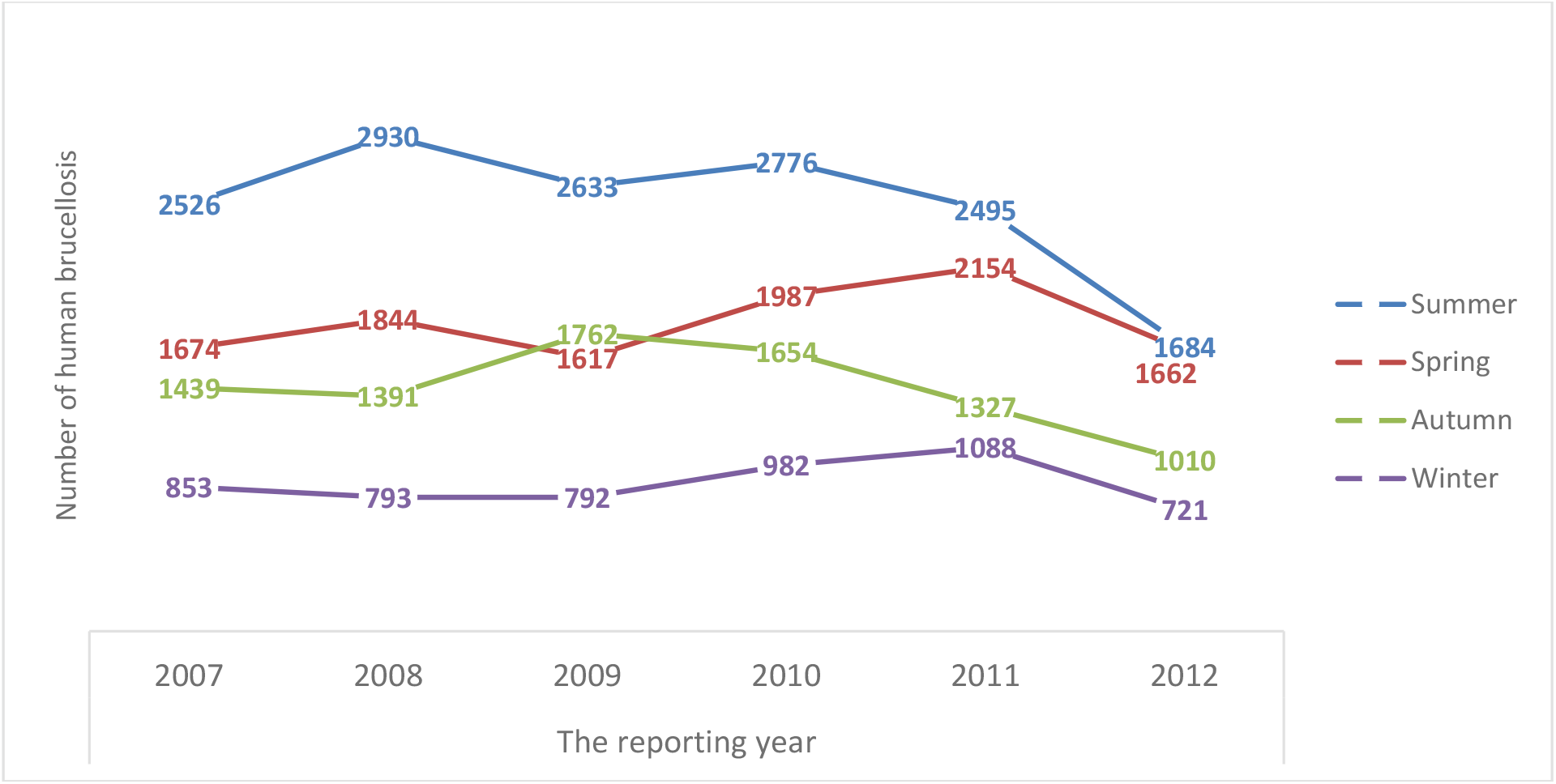
Seasonal distribution of Brucellosis cases in Iraq during 2007-2012.

**Figure 5:**
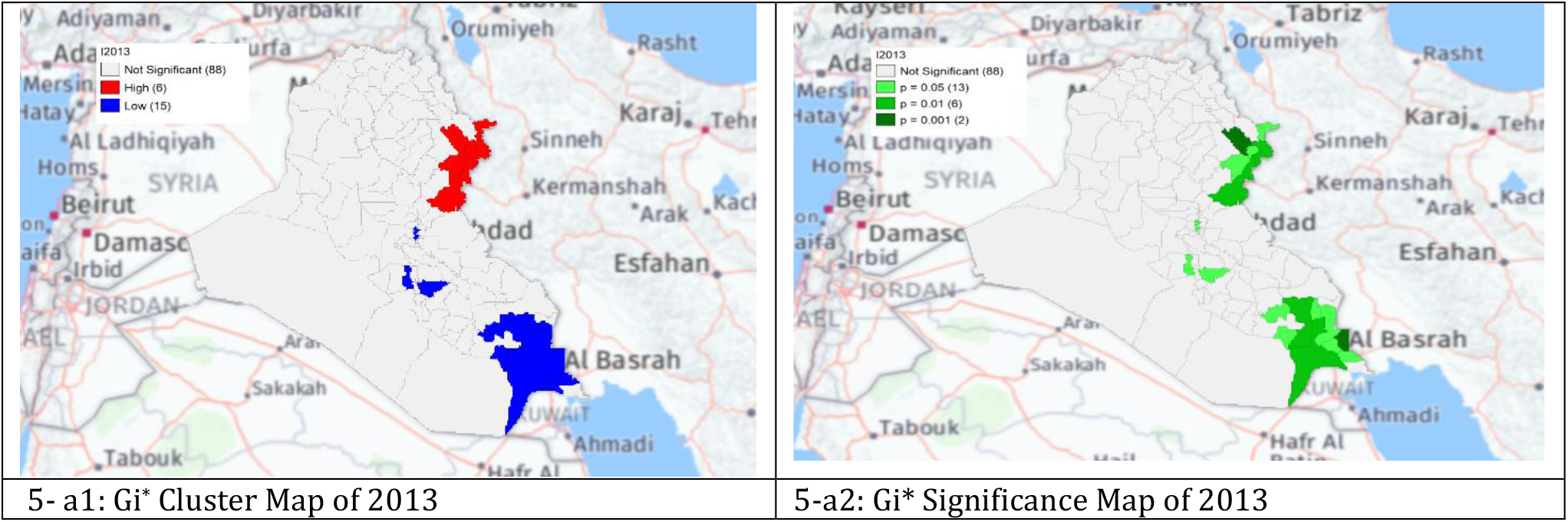
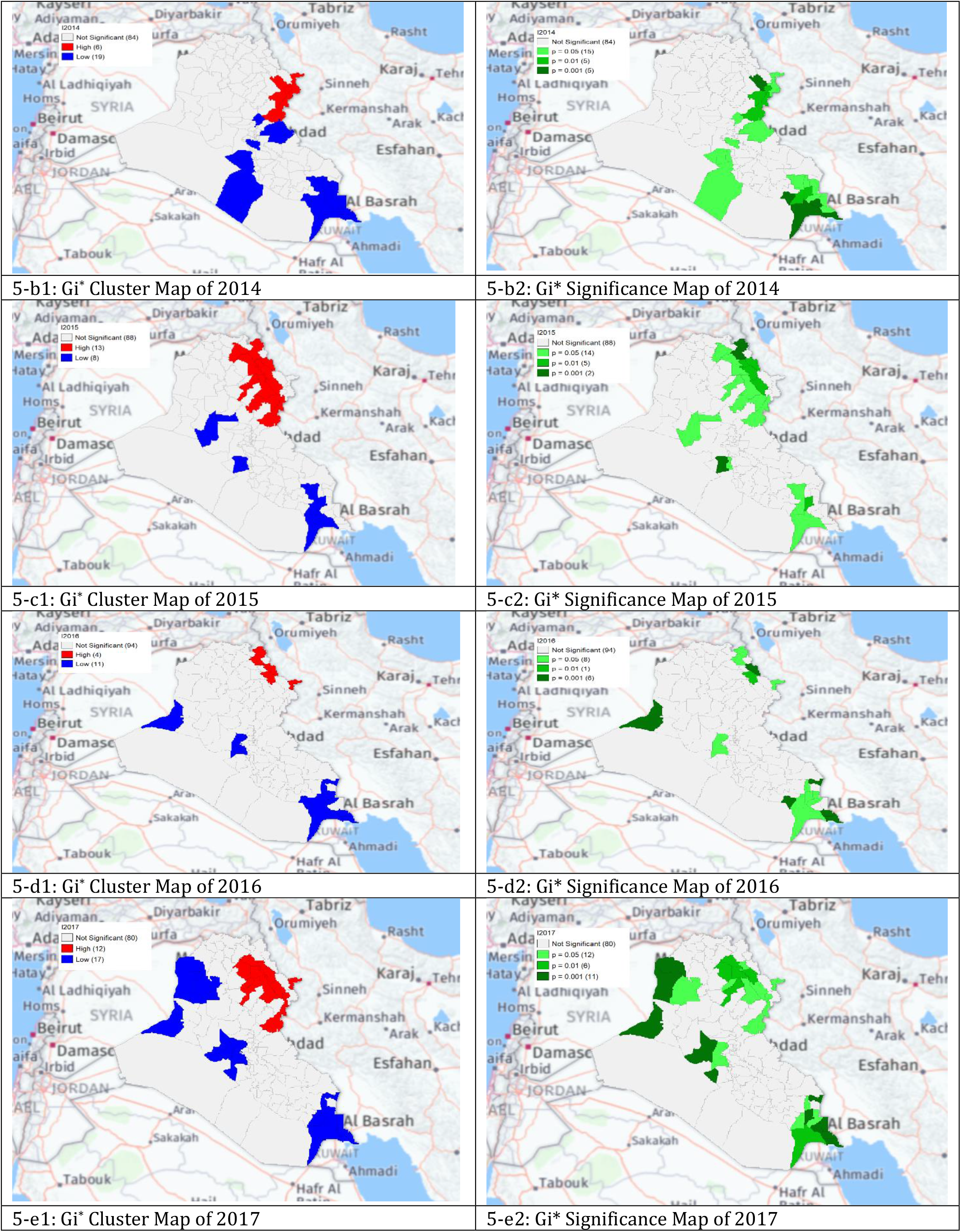
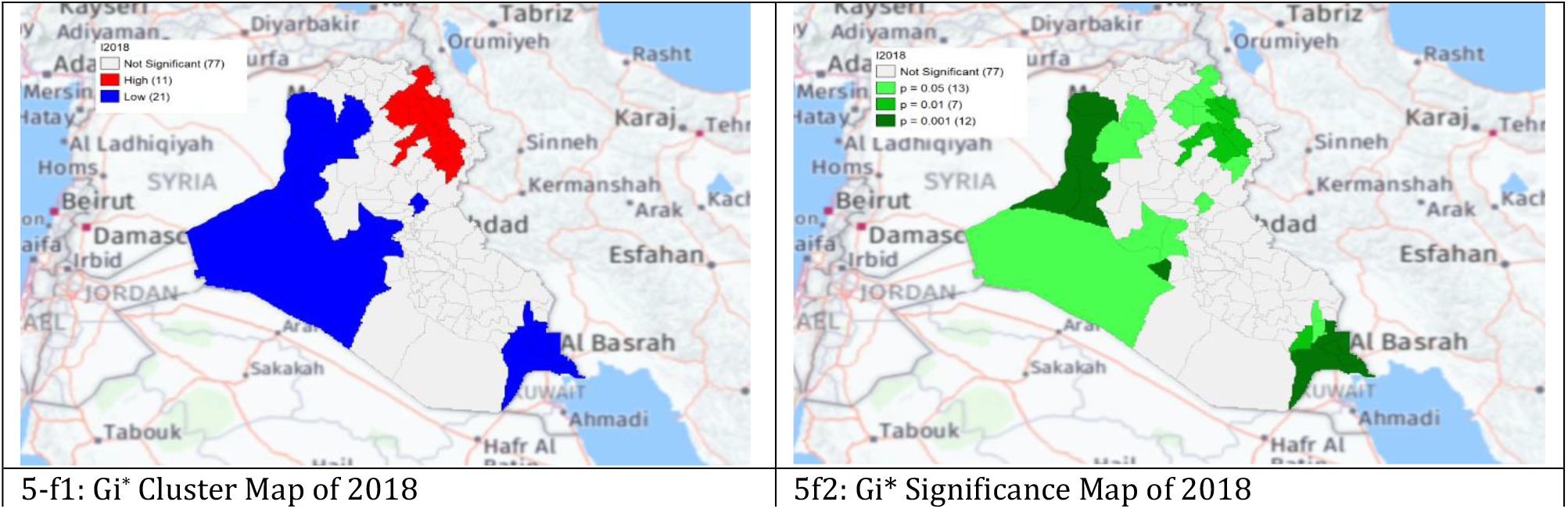
Gi* Cluster and Gi* Significance maps of human brucellosis cases in Iraq.

Most cases were females (58.02%), with an increasing frequency since 2016. Nearly 62% of the cases were 15-45 years, with no apparent change in the age groups affected throughout the years

Seasonal distribution of HB cases was constant throughout the year. Most of the cases occurred in summer, followed by spring.

The local Getis-ord Gi* statistics showed increased hot spots (High-High clusters) and cold spots (Low-Low clusters) from 2013 to 2018. Hot spots were located in the North and North-eastern parts, that is, in Choman, Soran, and Erbil (Erbil); Raniya (Sulaymaniyah); Akre (Nineveh); Kirkuk and Dibis (Kirkuk); Alshirqat, Baiji, and Tikrit (Salah Al-Din); and Amedi (Dahuk) In contrast, cold spots (Low-Low clusters) were located in districts located in provinces of Thiqar, Muthanna, Maysan, Anbar, Najaf, and Baghdad. District Dibis in Kirkuk shifted from a cold spot in 2015 to a hot spot in 2016. District Koisanjaq (Erbil) shifted from a hot spot in 2014 to a cold spot in 2015.

## Discussion

Analyzing the spatiotemporal distribution of HB is important in understanding the epidemiology of the disease in the country and helps forecast the future situation. The temporal trend revealed a decline in HB cases since 2012 that may be due to veterinary protective and control measures through vaccinating livestock that have been implemented since 2007. Although several vaccination rounds have been implemented since disruptions occurred during ISIL events that negatively affected the control program. ^5^,^6^ Nevertheless, animal health campaigns were relaunched soon after liberation in 2017.^7^

The number of females has been constantly higher than males, with increased reporting frequency since 2015. This can be due to increased exposure of women to infected animals and animal products through housekeeping and farming activities. ^5 **(11,12,13)**^ This contradicts what has been found in research from Germany^1^, Italy^8^, and Iran.^9^ Therefore, the potential differences should be investigated further to detect whether the difference is due to different proportions of males in different countries or biological differences that enhance contracting brucellosis. Nearly two-thirds of the patients were between 15 and 45 years old, which can negatively affect the economic status of the patients and their families and the quality of life for the patients.^10^ People in this age group may consume more locally made dairy products due to a lack of knowledge regarding the importance of pasteurized milk and milk products. However, this age category was very broad and could have been classified into two to three age groups to detect the most commonly affected age group. ^**(14-17)**^

Most HB cases were in the Northern provinces during the first period (2007-2012), especially Salah Al-Din, Sulaymaniyah, Nineveh, Erbil, Kirkuk, and Dahuk. Northern provinces share an extensive border with Iran, Turkey, and Syria and direct borders with other Iraq provinces, which, in turn, share borders with Jordan, Saudi Arabia, and Kuwait. ^11,12^ Sheep and goat husbandry has been practiced in the Northern region of Iraq since the earliest times due to the nature of these provinces, which is characterized by broad grass-covered terrain, undulating hills steep, and craggy mountains well suited to sheep and goat grazing. Livestock and its products dominate the area’s economy and the nutrition of its inhabitants.^13^ Therefore, the high clustering of brucellosis in this area may be due to area grazing strategy and animal population density, owner’s ignorance of the hazards of the disease, unhygienic disposal of infected animals as well as aborted fetuses or placental membranes, uncontrolled rigid restriction of the movement of diseased animals.^14^ The infected animals must be eradicated by slaughtering and burning because there is no curable medical therapy for animal brucellosis. In Iraq, these animals are expensive, so the farmers firmly refuse to slaughter and burn them. As a result, the infection will persist and be transmitted to their children by breastfeeding and other animals who consume their infected milk. On the other hand, humans may consume this infected milk unpasteurized, resulting in infection and areas endemic with brucellosis to animals and humans.^15^

The seasonal variation in the occurrence of human brucellosis in Iraq, that is, the decline in the number of HB cases in winter and the rise in spring and summer seasons, could be explained by high exposure to the disease during spring and summer when the deliveries of animals, increased milk production, and contamination occur. ^16,17^

The clustering of HB cases was in the North and North-Eastern region of Iraq, mainly in areas in the vicinity of Zagros mountains, which contain dense oak forests and fertile soil with a high density of sheep and goats; the main economic activity in the area is animal farming. The Zagros Mountains are also the route of seasonal migration for nomads. 18 Human-to-human transmission is rare. Clusters of human cases are most likely a result of animal processing, more intensive agricultural production zones, and similar socio-cultural practices.^8^

Transboundary transfer of animal brucellosis in the region from the neighboring countries such as Iran, Turkey, and Syria were provoked by war and political instability, lack of immunization and animal quarantine, frequent trading, low awareness and poor knowledge of HB prevention and control, residents with poor sanitary conditions easily exposed to Brucella contaminated food and water sources.

On the contrary, the low cluster areas may be due to having economic superiority, standardized industrial manufacturing for cow or sheep-related products, good sanitary habits, awareness of HB, and easy access to immediate treatment after infection.^4,19^ In addition, several changes in the surveillance case definition, the diagnostic methods, and the reporting of human brucellosis that occurred during the second period of the study could have also contributed to the low number reported compared to the first period. For example, the primary diagnostic method during the first period was the Rose-Bengal test, which has variable sensitivity and specificity based on the exposure history, stage of infection, and prior infection history. In contrast, the diagnostic test used during the second period was ELISA IgM, IgG, and culture. Although all suspected cases are initially reported to the surveillance section, their final classification as suspected or confirmed may vary by governorates based on their testing and interpretation capacity. ^20^Another example the data were collected in an aggregate format based on gender, age group, and province. In contrast, the data were collected in a case-based format during the second period of the study.^20^

This study has several strengths. First, this is the first study to include data from 10 years in a detailed spatiotemporal analysis. The consistency of the epidemiological characteristics across ten years can lead to public health interventions on where efforts should be spent. That is, the fact that the infection is consistently higher among females may direct health promotion activities to design educational materials that target females working in the animal husbandry industry on dealing with animals and their products and delivering them using personal protective measures. The second is using an innovative statistical approach to detect whether the clustering of cases is significant or random in addition to follow-up of the progress of clusters across seven years, which clearly showed a continued increase in the north and northeastern areas and continuous decline in the southern areas.

This study also has limitations. First, data regarding important risk factors of human brucellosis, such as occupation, rural or urban areas, comorbidities, and treatment protocol, were lacking. Had this information been available, it would have facilitated a better understanding of the epidemiology and characteristics of human brucellosis in Iraq.^20^

## Conclusions

Despite the declining incidence of HB in Iraq during 2013-2018, HB is still endemic and constitutes a public health problem. Most cases were reported in the summer and spring seasons among females and those aged 15-45 years. HB cases presented significant spatial clustering in northern and northeastern areas. Preventive measures such as health education activities should be performed in high-risk areas. Adopting the Quarantine-Slaughter-Immunization strategy and One Health Approach is crucial in controlling the disease. This can be achieved through multisectoral coordination and coordination with neighboring countries in the control programs.

## Data Availability

All data produced in the present study are available upon reasonable request to the authors

## Notes

### Competing Interest Statement

The authors have declared no competing interest.

### Funding Statement

This study did not receive any funding

### Author Declarations

Administrative and ethical approval was granted from the Public Health Directorate/Iraq MOH.

